# Proper Name Recall as an Early Indicator of Preclinical Alzheimer’s Disease Pathology

**DOI:** 10.1101/2025.06.05.25329051

**Authors:** Kimberly D. Mueller, Anja Soldan, Rebecca Langhough, Davide Bruno, Ainara Jauregi-Zinkunegi, Kristin Basche, Madeline Hale, Deling He, Abhay Moghekar, Bruce Hermann, Marilyn Albert, Corinne Pettigrew

## Abstract

**Background:** Early detection of Alzheimer’s disease (AD) is crucial; however, standard neuropsychological tests often lack sensitivity. Process scores, such as proper name (PN) recall from Logical Memory (LM), may improve the detection of AD-related biomarker positivity. We examined whether baseline PN recall predicted future cerebrospinal fluid (CSF) amyloid (Aβ42/Aβ40) and tau (pTau_181_) status, and whether biomarker status predicted PN recall trajectories.

**Methods:** We analyzed 271 cognitively unimpaired BIOCARD participants (mean age = 57.3, 60.3% female, mean follow-up = 15.5) using logistic regression and mixed-effects models to examine the associations between PN recall and CSF biomarkers.

**Results:** Higher baseline PN recall predicted lower amyloid positivity (odds ratio [OR] = 0.72, p = 0.015). Amyloid and tau positivity have been linked to a faster decline in PN. Biomarker-positive participants in the biomarker-negative group lacked practice effects.

**Conclusions:** PN recall predicts future AD biomarker positivity and may enhance early detection of AD-related cognitive decline.

## 1. BACKGROUND

Identifying early cognitive decline associated with Alzheimer’s disease (AD) biomarkers is crucial for targeting individuals who may benefit the most from early interventions in the preclinical stage of the disease.^1^ However, neuropsychological tests commonly used in longitudinal AD studies often lack sensitivity to the earliest pathological changes, making early detection challenging^2, 3^. Since many cohort studies and clinical trials have historically relied on these measures for follow-up, re-scoring existing tests could offer cost-effective and time-efficient solutions while reducing participant burden.

Cognitive “process scores,” the assessment of strategies or types of responses that differ from a traditional total score^4^, are a promising alternative. For example, using the Logical Memory (LM) story recall task from the Wechsler Memory Scale-Revised (WMS-R),^5^ our group examined process scores based on the “lexical categories” (i.e., verbs and numerical expressions) of recalled words. Particularly, “proper names” is a lexical category referring to the names of people or places (e.g., *Joe, New York*). They are particularly challenging to encode and retrieve from memory, above and beyond that of common nouns^6^, even when considering cognitive reserve.^7^ This may be attributed to the limited descriptive meaning, adherence to unique morpho-syntactic rules,^8^ and the involvement of fewer neural networks in retrieval compared to common nouns or verbs.^9^ Additionally, proper name retrieval engages brain regions vulnerable to AD, such as the left anterior temporal lobe and parahippocampal gyri, making proper name recall a potentially sensitive early marker of AD-related cognitive decline.^6, 9^

Indeed, using LM data from cognitively unimpaired participants from the Wisconsin Registry for Alzheimer’s Prevention (WRAP), we found that baseline proper name delayed recall, but not traditional total scores, were cross-sectionally associated with amyloid-PET deposition.^10^ While these data suggest that proper name retrieval may be sensitive to AD biomarkers among cognitively unimpaired participants. These results have not yet been replicated and validated in other cohorts.

This study aimed to replicate and extend previous WRAP findings on proper name recall using the Biomarkers of Cognitive Decline Among Normal Individuals (BIOCARD) cohort. Our objective was to: **Aim 1:** Test whether baseline delayed proper name recall in cognitively unimpaired adults predicts the most recent cerebrospinal fluid (CSF) amyloid and p-tau positivity; **Aim 1a**: Confirm that only delayed—not immediate—recall is linked to biomarker status, suggesting retrieval or consolidation deficits; **Aim 2**: To examine whether biomarker status predicts longitudinal decline in proper name recall, and **2a**: whether these effects are stronger than the total score decline. **Aim 3**: Assess whether biomarker status moderates practice effects—repeated test exposure benefits—on story recall trajectories.

We hypothesized that (1) baseline delayed proper names would predict CSF Aβ42/Aβ40 and pTau181 status; (2) associations would be stronger for proper names than total scores; (3) biomarkers would be related to a decline in delayed, but not immediate, recall; (4) proper name models would yield larger effect sizes; and (5) biomarker-positive individuals would show reduced practice-related gains over time.

## 2. METHODS

### 2.1 Standard Protocol Approvals, Registrations, and Patient Consents

The BIOCARD study was approved by Johns Hopkins Medical Institutional Review Board. Written informed consent was obtained from all participants.

### 2.2 Study Design and Participants

Data were drawn from the BIOCARD study, a longitudinal cohort study designed to identify predictors of cognitive decline and dementia in initially cognitively unimpaired individuals. Established at the National Institutes of Health in 1995, 349 participants were recruited between 1995 and 2005 through the Geriatric Psychiatry Branch at NIMH. Annual cognitive and clinical assessments, along with biennial CSF, blood, and MRI collections, were conducted.^11^ The study paused in 2005 and resumed at Johns Hopkins University in 2009, with biennial CSF and MRI collection restarting in 2015.

Participants were assigned consensus clinical diagnoses (normal, MCI, impaired not MCI, or dementia) based on the NIA-AA criteria, blinded to biomarker and APOE-ε4 status.^12, 13^ This study included 271 participants who had at least one CSF biomarker measurement, item-level LM Story A data from two or more visits, and were cognitively unimpaired at baseline. The exclusion criteria included participants who withdrew or had not re-enrolled (n=25) and those classified as amyloid-negative/tau-positive at their most recent CSF visit (n=28), as these individuals may represent non-Alzheimer’s pathological change^14, 15^.

### 2.3 Cognitive Outcome Measures from LM Story A

Participants completed an annual cognitive assessment that included LM story A (immediate and delayed recall), a part of the WMS-R.^5, 11^ This subtest evaluates episodic memory by asking participants to immediately retell a short story after hearing it. They were then asked to remember it for a delayed recall 20-30 minutes later. If needed, a brief prompt (e.g., “The story was about a woman who was robbed”) was provided.

#### 2.3.1 Total Score (traditional scoring procedure)

LM Story A consisted of 25 distinct idea units (e.g., names, locations, and actions). Examiners marked each expressed unit and noted nonverbal responses for scoring. The manual allows flexibility for accurate paraphrasing, except for proper names, which require near-verbal recall. Each correct unit scored 1, and missed units scored 0. Immediate and delayed recalls were scored separately, with each score ranging from 0 to 25.

#### 2.3.2 Proper Name Score (novel “process score”)

Unlike traditional scoring procedures, process scores capture the qualitative nature of behavior. ^4, 16^ Using de-identified BIOCARD data managed in REDCap at the University of Wisconsin–Madison, ^17^ item-level responses were transcribed and processed with part-of-speech tagging to identify four proper names in Story A. Immediate and delayed proper name scores (0–4 each) were derived. The focus was on delayed retrieval and its association with biomarkers, although immediate scores were also examined for comparison purposes.

### 2.4 CSF Measures

CSF samples were collected via lumbar puncture after an overnight fast, while the study was conducted at the NIH (1995–2005) and JHU (since 2015). As described previously^18^, the samples were aliquoted into polypropylene cryotubes that were kept on dry ice and immediately transferred to a -80°C freezer for long-term storage. Samples were thawed for the first time after collection to measure Aβ_40_, Aβ_42_, t-tau, and pTau_181_ levels using fully automated electrochemiluminescence assays on the Lumipulse G1200 platform (Fujirebio Diagnostics, Inc.).

We used each participant’s most recent CSF sample (mean [SD] = 10.7 [7] years from baseline) to maximize the detection of biomarker positivity, as most participants were middle-aged at baseline. CSF Aβ42/Aβ40 and pTau181 were dichotomized using Lumipulse cut points (Aβ42/Aβ40 ≤ 0.070; pTau181 ≥ 51.0) to define separate amyloid (A) and tau (T) status groups.^19^ Primary analyses examined A and T positivity separately; secondary analyses assessed combined A/T groups (A−T−, A+T−, A+T+) per NIA-AA criteria. ^20, 21^ Sensitivity analyses used log-transformed continuous biomarker values to account for skewness.

### 2.5 APOE Genetic Status

*APOE*-ε4 genetic status was determined by restriction endonuclease digestion of PCR-amplified genomic DNA (Athena Diagnostics, Worcester, MA, USA). Binary indicators for *APOE*-ε4 genetic status were created (*APOE*-ε4 carriers of at least one allele = 1, ε4 non-carriers = 0).

### 2.6 Practice Effects

To assess whether repeated exposure to LM Story A influenced scores or moderated the relationship between CSF biomarkers and memory trajectories, practice was defined as the number of prior test exposures (total visits minus one). This approach has previously accounted for the variability in WRAP longitudinal data.^22^

### 2.7 Statistical Analyses

#### Aim 1

We used logistic regression to test whether baseline PN-delayed predicted CSF biomarker positivity at each participant’s most recent CSF assessment. Outcomes included binary amyloid (Aβ42/Aβ40; A+/−) and pTau181 (T+/−) status. As a secondary outcome, we examined a binary A/T variable (A-/T-vs. A+/T+) to represent individuals who were least and most likely to show subtle cognitive impairment. For each biomarker outcome, we ran three models: (1) proper names--unadjusted; (2) adjusted for baseline age, sex, education, and time from baseline to CSF collection; and (3) Model 2 + APOE-ε4 status.

#### Aim 1a

To compare proper name delayed (0–4) with immediate (0–4) and total (0–25) scores, we repeated the logistic models using these alternative LM predictors. Model performance was compared using Akaike’s Information Criterion (AIC), odds ratios (OR), and 95% confidence intervals (CI).

#### Aims 2 and 2a

We used linear mixed-effects models to examine the association between CSF biomarker status (A+/- or T+/-) and change in story recall from baseline to the most recent cognitive visit. The separate models tested the interactions between biomarker status and time for delayed proper names and total scores. Secondary models included immediate recall outcomes and a three-level A/T status (A-/T-, A+/T-, and A+/T+). The outcomes were standardized (M=0, SD=1). Time was modeled as age at visit (centered at 68 years) with a quadratic term. All models included random intercepts and slopes and were adjusted for sex, education, practice effects, age at CSF collection, biomarker group, and interactions between biomarker group and age/age². Nonsignificant CSF group×age² interactions were excluded.

Sensitivity analyses used log-transformed continuous CSF Aβ42/Aβ40 and pTau181 values, instead of categorical predictors. Secondary analyses examined longitudinal changes in immediate recall scores.

#### Aim 2a

To compare effect sizes across recall predictors, we computed standardized estimates and 95% CIs for all mixed models.

#### Aim 3

To test whether practice effects moderated biomarker–recall trajectories, we included three-way interactions (biomarker ×age×practice) in linear mixed models. We evaluated model fit using AIC, R², and variance inflation factors (VIFs). The final models retained random intercepts, linear age slopes, and covariates from Aim 2.

#### Additional Analyses

Demographic and clinical characteristics (Table 1) were compared between the A/T status groups using t-tests (continuous), chi-square (binary), or Kruskal-Wallis tests (non-normal variables). A significance level of p<.05 was used (uncorrected). All analyses were conducted using R 4.4.2.

## 3. RESULTS

Baseline demographic and clinical characteristics of 271 participants stratified by CSF A/T status at their most recent visit are presented in **Table 1**. Participants were 56.5 (SD=10.2) years at their baseline cognitive testing visit and underwent 15.3 (SD=6.7) years of story recall follow-up; participants who were A+/T- (n=34, mean age = 57.6, SD=6.4 or A+/T+ (n=63, age = 61.1, SD = 9) at their last CSF measurement were significantly older at baseline than those who were A-/T- (n=154, age = 53.6, SD = 10.5). The biomarker-positive group had a higher percentage of *APOE*-ε4 carriers than the biomarker-negative group (A-/T- =17%, A+/T- = 54%, and A+/T+ = 62%). The biomarker groups did not differ by years of education, sex, race, baseline MMSE, or the time point of the last CSF collection, which was an average of 10.7 years (SD = 7.4) after the baseline cognitive testing visit.

### 3.1 Baseline LM scores and most recent CSF biomarker Status (Aims 1 & 1a)

#### Proper name delayed

Delayed proper name recall was linked to lower odds of A+ status across all models (Model 1 OR = 0.73, Model 2 OR = 0.69, Model 3 OR = 0.72, all p < 0.05). Similar associations were found for T+ status in the unadjusted and demographics-adjusted models (Ors = 0.76, p = 0.02), although the fully adjusted model was not significant (OR = 0.78, p = 0.085; Table 2).

#### Total score delayed

Logistic regression (Supplemental Table 1) showed that higher delayed total scores were associated with lower odds of A+ in the unadjusted and demographics-only models (ORs = 0.90, p ≤ 0.02), but not in the fully adjusted model (p = 0.084). Similar patterns were observed for T+ status, with significance in the first two models (ORs = 0.89–0.90, p ≤ 0.04), but not in the fully adjusted model (p = 0.118). The ROC curves and AUCs for total and proper name delayed scores are shown in **Supplemental Figure 1**.

Secondary analyses for A/T two-level status (A-/T- vs. A+/T+ to fit the binary logistic regression) for proper name delay were nearly identical to the T+ models; higher delayed proper name recall was associated with lower odds of A+/T+ (Model 1: OR = 0.79, CI 0.64-0.98, p=0.031; Model 2: OR = .75, CI = .59 - .95, p = 0.017), but the relationship was nonsignificant in the *APOE* adjusted model. The delayed total score was not a significant predictor of A/T status in any model. In contrast, neither baseline proper name nor total score immediate was associated with any of the biomarker outcomes (i.e., A+/-, T+/-, all p>0.05, **Supplemental Tables 2 and 3**).

### 3.2 Longitudinal proper name and total score trajectories (Aims 2 and 2a)

#### Proper name trajectories

Linear mixed models (**Supplemental Table 4**) showed significant interactions between biomarker status and quadratic age. A+ and T+ groups declined faster in both delayed and immediate proper names than their negative counterparts (all p < 0.001; **Figure 2**). Practice effects were significant for delayed recall (β = 0.01, p = 0.02) but not immediate. Women and those with more education generally performed better on LM measures.

**Figure 1.**
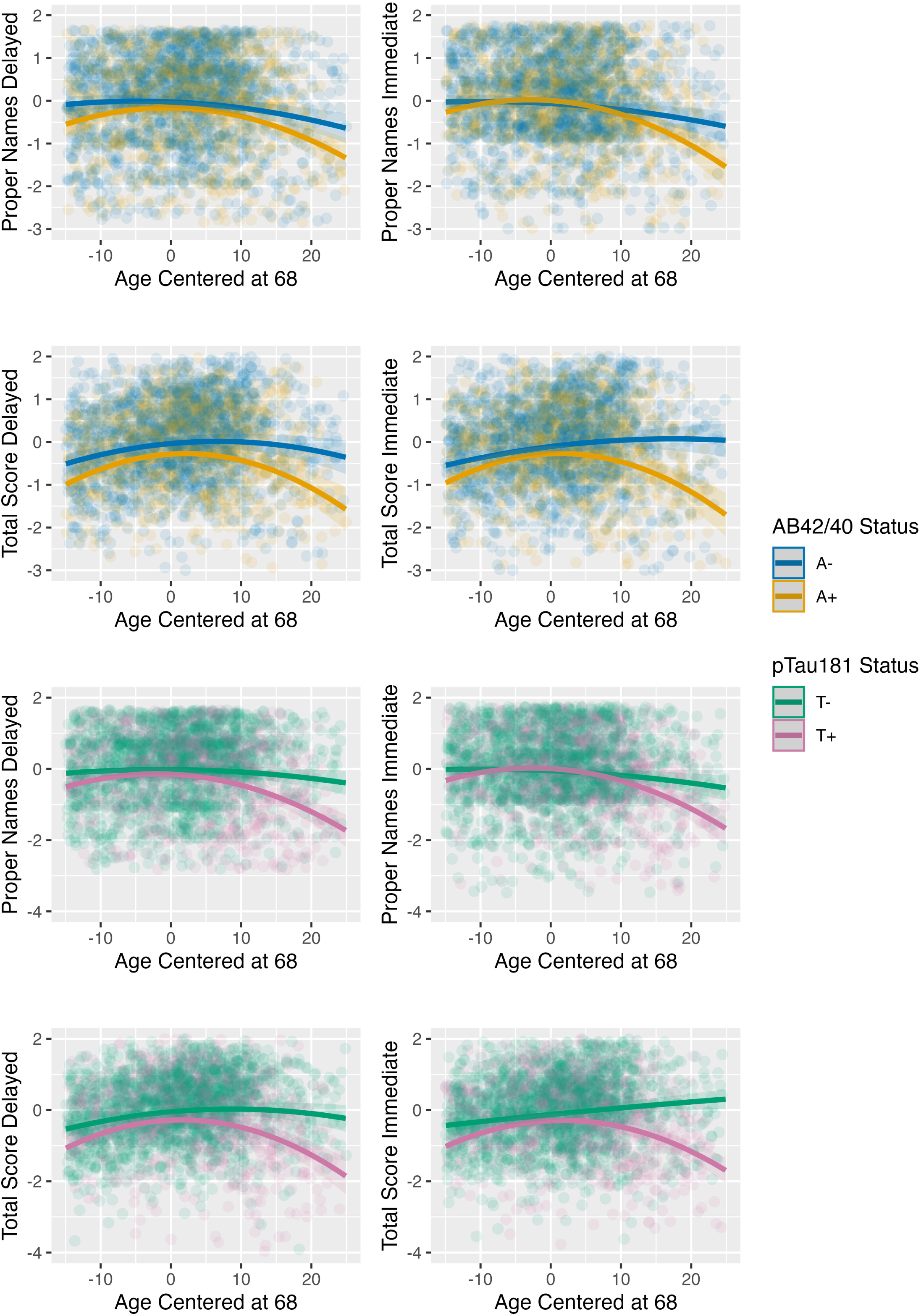
Amyloid (top) and Tau (bottom) Status Predicting Longitudinal Proper Names and Total Score Trajectories, Delayed (left) and Immediate Recall (right) from LM. Linear mixed-effects models including random effects for intercept and slope, age (centered), and a quadratic term for age (centered), co-varied for sex, education, practice effects, and age at CSF collection. Blue solid lines depict simple slopes and confidence intervals for amyloid-negative (A-); orange lines depict amyloid-positive (A+); green lines depict pTau_181_ negative (T-); magenta lines depict pTau positive (T+) groups. LM trajectories are from baseline cognitive testing to the last (most recent) visit (mean, SD years of follow-up = 15, 6); amyloid and tau status are from the last (most recent) CSF visit (mean years, SD, from last cognitive testing = 4.6, 0.4). n=271 participants.

**Figure 2.**
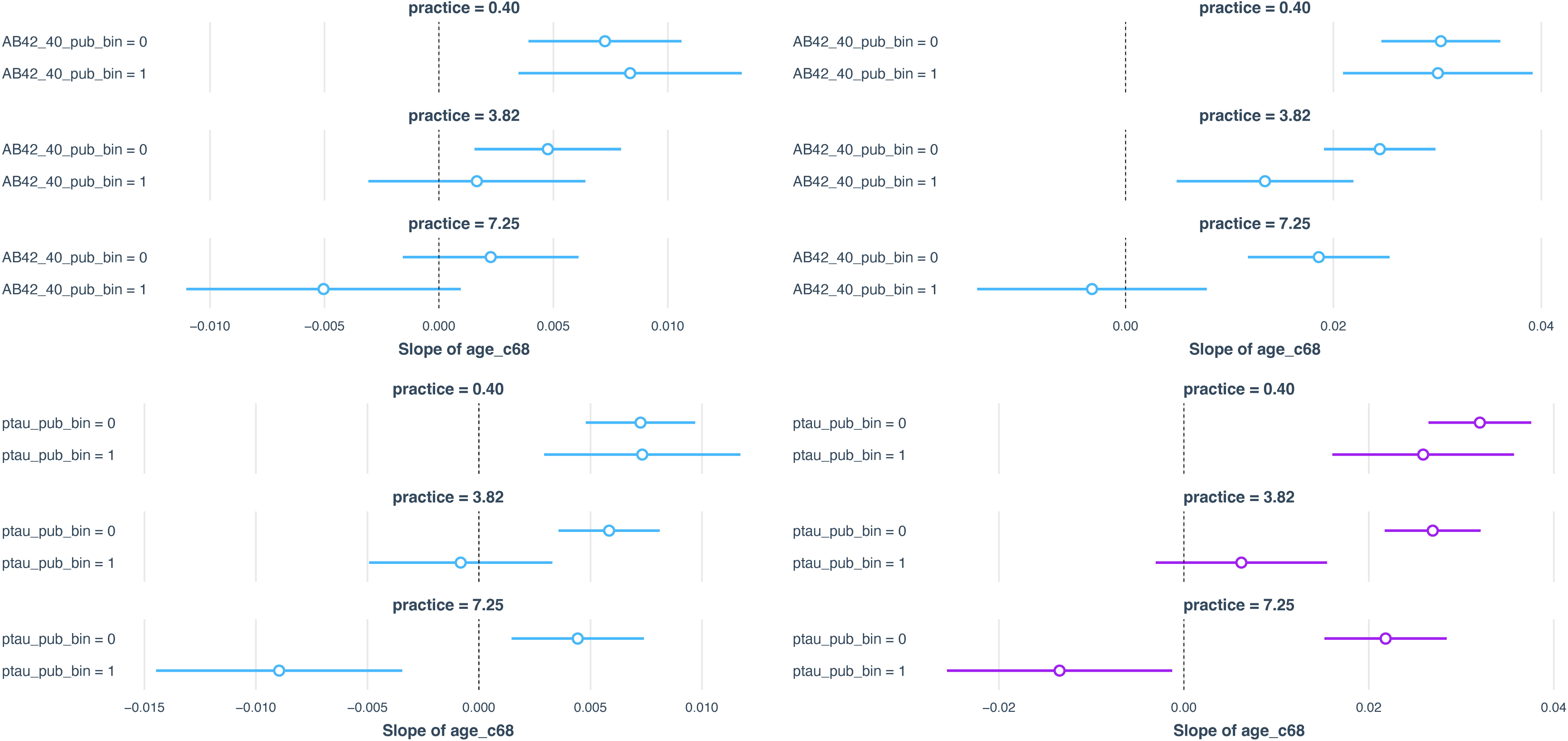
Proper name (left) and total score (right) simple slopes by amyloid (top) and tau (bottom) status, varied by levels of practice. The top panel represents the simple slopes and standard errors for proper names by Aβ_42/40_ status over levels of practice for n=271 participants. Time-varying practice is defined as participant’s LM visit number – 1 to represent the number of previous exposures to the test.^22^ Levels of practice from top to bottom are the mean – 1SD, mean, and mean + 1SD of prior exposures. The bottom panel shows the slopes and standard errors for p-Tau status. Standard error bars that did not cross 0 are significant at p<.05.

#### Total score trajectories

A+ and T+ groups showed significantly faster curvilinear decline in delayed total score recall compared to A− and T− groups (all p < 0.01). Similar patterns were observed for immediate recall (**Supplemental Table 5,**, **Figure 1**).

In both amyloid and tau models, greater exposure to the LM test was linked to higher delayed total recall scores (β = 0.01, p = 0.02), but not immediate recall. Females outperformed males in all total recall measures, while education was only associated with better total and PN delayed recall. Standardized coefficients (**Table 3**) showed the largest effect for T+ status × age² in the total immediate recall model (β = -0.29), followed by similar interactions in delayed total (β = -0.27), immediate total (A+; β = -0.25), and proper name immediate recall (T+; β = -0.22).

#### Sensitivity analyses

Using 3-level A/T status (A−/T−, A+/T−, A+/T+), A+/T+ individuals showed faster decline in delayed proper name and total recall, mirroring results from binary models (**Supplemental Table 6**); similar significant patterns were found for continuous log-transformed pTau181 and Aβ42/Aβ40 levels.

### 3.3 Practice effects (Aim 3)

Three-way interactions among biomarker status, age, and practice revealed that A+ and T+ individuals showed reduced benefit from repeated testing on delayed proper names (A+ β = - 0.001, p = 0.009; T+ β = -0.002, p < 0.0001), with steeper declines compared to A−/T− groups. Similar effects were observed for immediate recall in the T+ model (β = -0.004, p = 0.002) but not in the A+ model. For total score delayed, three-way interactions were significant across all models (A+ β = -0.003 to -0.004, p ≤ 0.001; T+ β = -0.004 to -0.005, p < 0.0001), indicating that the A+/T+ group showed less practice-related improvement over time (**Figure 2; Supplemental Table 7).**

## 4. DISCUSSION

In this study, we examined the association between a novel metric of lexical-semantic processing, recall of proper names from the LM story recall task, and CSF amyloid and pTau biomarkers of AD pathology in cognitively unimpaired and largely middle-aged participants at baseline. Key findings include: (1) baseline delayed proper name recall from Story A was associated with CSF amyloid and pTau positivity collected an average of 10 years later; (2) CSF AD biomarker positivity was associated with a faster decline in both proper name and total score recall trajectories over an average of 15 years; and (3) practice effects, reflecting the number of prior exposures to the LM task, were moderated by biomarker status on proper name delayed recall, with amyloid and tau-positive groups deriving less benefit from repeated testing. Taken together, these results provide additional insights into possible lexical-semantic processing deficits in preclinical AD and suggest that cognitive process scores from longitudinal neuropsychological tests may enhance sensitivity to early cognitive decline in AD.

This study builds on prior WRAP findings ^10^ on [^11^C]PiB PET amyloid biomarkers and validated them in the BIOCARD cohort using CSF amyloid and pTau biomarkers. Each additional proper name recalled during delayed recall was associated with a 31% reduced risk of amyloid positivity and a 26% reduced risk of pTau positivity, whereas delayed total score recall measures were linked to a 10% reduced risk of biomarker positivity. These findings, using Story A alone (range = 0-4), are consistent with those from WRAP, which combined stories A and B (range = 0-9).^23^ Notably, only baseline delayed proper name recall, but not immediate recall, predicted biomarker positivity, confirming our hypothesis and suggesting an early lexical retrieval deficit rather than encoding impairment.

The findings were somewhat weaker for the CSF tau (T+/−) models and combined amyloid/tau (A/T) models than for the amyloid-only (A+/−) models. This indicates that proper name recall in cognitively unimpaired individuals may be more closely linked to future amyloid status than to tau status. Prior research suggests that CSF Aβ42/Aβ40 is a stronger predictor of cognitive decline than pTau181 when considered separately, possibly because it reflects both AD-specific and mixed-pathology processes.^24,25^ Importantly, proper name recall was more predictive of future biomarker positivity than the traditional total recall score, highlighting its potential as a brief, accessible tool to identify at-risk individuals, potentially reducing the need for more invasive and expensive testing.

Our longitudinal findings further reinforced the link between proper name recall and preclinical AD pathology. Longitudinal models showed that biomarker-positive status was linked to a faster decline in both proper name and total delayed recall score, extending prior WRAP findings. While effect sizes were modest and similar across outcomes, pTau_181_ had a slightly stronger effect on total score decline (β = 0.27) than on proper name trajectories (β = 0.18), possibly because of the narrow range for proper names (0–4). The baseline proper name scores were already low for some participants, limiting the observable decline. Nevertheless, the sensitivity of this brief measure supports its utility in cognitive assessments, offering a more efficient alternative to longer recall tasks.

Baseline immediate proper name and total score measures did not predict future biomarker status, which was consistent with our hypothesis. However, biomarker positivity was significantly associated with proper name and total score immediate decline over time, and both pTau and amyloid positivity showed larger effect sizes for immediate decline than for delayed decline. These findings suggest that encoding difficulties increase over time in preclinical AD and affect delayed recall. The lack of practice effects on immediate memory but their presence in delayed recall further supports the evolving encoding deficits in preclinical AD^22, 26, 27^. These results are consistent with a longitudinal study among clinically normal adults, which showed declines and diminished practice effects on immediate measures of memory retrieval in response to emerging AD pathology, as measured by very early Aβ accumulation (< Centiloid [CL] 20 or CL 40).^28^ Our study follows this pattern: we observed that biomarker-positive individuals exhibited *diminished* practice effects on the LM task compared to biomarker-negative peers. In other words, cognitively normal adults without AD pathology tend to improve or maintain their performance upon repeated testing, whereas those with underlying amyloid or tau show little to no improvement over time. This difference in practice-related gains implies that subtle cognitive decline may already be underway in the biomarker-positive group, even though they remain clinically unimpaired. Such attenuation of practice effects in the presence of AD biomarkers aligns with other research showing that asymptomatic amyloid-positive individuals fail to show normal improvements in serial cognitive testing^22, 26, 27^. Diminished practice effects may reflect early A-related reductions in learning, whereas the delayed recall and retrieval deficits observed in more advanced stages of preclinical AD pathology may be more strongly associated with abnormal tau deposition in the medial temporal lobe.^28–30^ Future studies combining measures of amyloid and tau PET with story recall measures are needed to address this question directly.

Proper names are linguistically and cognitively demanding; they lack semantic richness, have no synonyms, and often appear arbitrary, increasing the retrieval burden. ^31–33^ Their phonological diversity also complicates recall.^34^ These features may explain why proper name recall is particularly vulnerable in preclinical AD. Prior studies have mostly used isolated name-face or place-name tasks; in contrast, our use of narrative-based name retrieval, a more naturalistic form, has received limited experimental attention. This may tap into episodic memory networks,^35^ and may also rely on emotional, social, or cultural aspects of memory.^36^ Future work should explore the mechanisms by which early Aβ and tau pathology disrupt proper name encoding and retrieval, with the potential to refine early detection tools.

A strength of this study is the replication of a novel lexical-semantic measure using a widely used neuropsychological test in a well-characterized longitudinal cohort. We showed associations between CSF biomarkers and proper name recall over 15 years and found attenuated practice effects in biomarker-positive participants, highlighting the importance of modeling practice as a latent factor in longitudinal cognitive analyses.

However, limitations include a sample enriched for parental history of AD, high education, and limited racial/ethnic diversity, reducing generalizability. Proper name familiarity may also vary culturally and linguistically, thereby influencing performance. Additionally, the narrow scoring range for proper names likely contributed to the small effect sizes, which, while statistically significant, may not reflect meaningful clinical differences. Further studies are needed to validate these findings in a broader and more representative sample.

In conclusion, proper name recall is a sensitive marker of early AD pathology, providing insight into lexical-semantic memory changes in preclinical stages. As name retrieval is crucial for social interactions, impairments may contribute to isolation and downstream cognitive decline. Understanding the neural and behavioral mechanisms of name retrieval difficulties could inform efficient screening tools for early AD detection and support the development of non-pharmacological interventions for individuals with cognitive impairment and dementia.

## Data Availability

Anonymized data used in the analyses presented in this report are available on request from qualified investigators.

https://www.biocard-se.org

## Acknowledgments

The authors thank the members of the BIOCARD Scientific Advisory Board who provided continued oversight and guidance regarding the conduct of the study, including Drs. John Csernansky, David Holtzman, David Knopman, Walter Kukull, and Kevin Grimm, and Drs. John Hsiao and Laurie Ryan, who provided an overview of the National Institute on Aging. The authors thank the members of the BIOCARD Resource Allocation Committee who provided ongoing guidance regarding the use of the biospecimens collected as part of the study, including Drs. Constantine Lyketsos, Carlos Pardo, Gerard Schellenberg, Leslie Shaw, Madhav Thambisetty, and John Trojanowski. The authors acknowledge the contributions of the Geriatric Psychiatry Branch of the intramural program of the NIMH, which initiated the study (principal investigator: Dr. Trey Sunderland). The authors also thank Dr. Karen Putnam, who has provided ongoing documentation of the Geriatric Psychiatry Branch study procedures and data files received from NIMH.

## Consent Statement

The BIOCARD study was approved by the Johns Hopkins Medical Institutional Review Board, and all procedures were performed in accordance with the ethical standards laid down in the 1964 Declaration of Helsinki and its later amendments. Written informed consent was obtained from all participants.

## REFERENCES

1. Rafii MS, Aisen PS. Detection and treatment of Alzheimer’s disease in its preclinical stage. Nature aging 2023;3:520–531.

2. Rentz DM, Parra Rodriguez MA, Amariglio R, Stern Y, Sperling R, Ferris S. Promising developments in neuropsychological approaches for the detection of preclinical Alzheimer’s disease: a selective review. Alzheimer’s research & therapy 2013;5:1–10.

3. Aizenstein HJ, Nebes RD, Saxton JA, et al. Frequent amyloid deposition without significant cognitive impairment among the elderly. Archives of neurology 2008;65:1509–1517.

4. Milberg WP, Hebben N, Kaplan E, Grant I, Adams K. The Boston process approach to neuropsychological assessment. Neuropsychological assessment of neuropsychiatric and neuromedical disorders 2009;3:42–65.

5. Wechsler D. Wechsler memory scale-revised. Psychological Corporation 1987.

6. O’Rourke T, de Diego Balaguer R. Names and their meanings: A dual-process account of proper-name encoding and retrieval. Neuroscience & Biobehavioral Reviews 2020;108:308–321.

7. Montemurro S, Mondini S, Crovace C, Jarema G. Cognitive reserve and its effect in older adults on retrieval of proper names, logo names and common nouns. Frontiers in Communication 2019;4:14.

8. Langendonck WV. Berlin, New York: De Gruyter Mouton, 2007.

9. Semenza C. Retrieval pathways for common and proper names. Cortex 2006;42:884–891.

10. Mueller KD, Koscik RL, Du L, et al. Proper names from story recall are associated with beta-amyloid in cognitively unimpaired adults at risk for Alzheimer’s disease. Cortex 2020;131:137–150.

11. Albert M, Soldan A, Gottesman R, et al. Cognitive changes preceding clinical symptom onset of mild cognitive impairment and relationship to ApoE genotype. Current Alzheimer Research 2014;11:773–784.

12. Albert MS, DeKosky ST, Dickson D, et al. The diagnosis of mild cognitive impairment due to Alzheimer’s disease: recommendations from the National Institute on Aging-Alzheimer’s Association workgroups on diagnostic guidelines for Alzheimer’s disease. Focus 2013;11:96–106.

13. McKhann GM, Knopman DS, Chertkow H, et al. The diagnosis of dementia due to Alzheimer’s disease: Recommendations from the National Institute on Aging-Alzheimer’s Association workgroups on diagnostic guidelines for Alzheimer’s disease. Alzheimer’s & dementia 2011;7:263–269.

14. Jack Jr. CR, Andrews JS, Beach TG, et al. Revised criteria for diagnosis and staging of Alzheimer’s disease: Alzheimer’s Association Workgroup. Alzheimer’s & Dementia 2024;20:5143–5169.

15. Jack Jr CR, Knopman DS, Chételat G, et al. Suspected non-Alzheimer disease pathophysiology—concept and controversy. Nature Reviews Neurology 2016;12:117–124.

16. Bruno D, Jauregi Zinkunegi A, Kollmorgen G, et al. A comparison of diagnostic performance of word-list and story recall tests for biomarker-determined Alzheimer’s disease. Journal of Clinical and Experimental Neuropsychology 2023;45:763–769.

17. Harris PA, Taylor R, Thielke R, Payne J, Gonzalez N, Conde JG. Research electronic data capture (REDCap)—A metadata-driven methodology and workflow process for providing translational research informatics support. Journal of Biomedical Informatics 2009;42:377–381.

18. Pettigrew C, Soldan A, Wang J, et al. Longitudinal CSF Alzheimer’s disease biomarker changes from middle age to late adulthood. Alzheimer’s & Dementia: Diagnosis, Assessment & Disease Monitoring 2022;14:e12374.

19. Dakterzada F, López-Ortega R, Arias A, et al. Assessment of the concordance and diagnostic accuracy between Elecsys and Lumipulse fully automated platforms and Innotest. Frontiers in Aging Neuroscience 2021;13:604119.

20. Jack Jr CR, Bennett DA, Blennow K, et al. NIA AA research framework: toward a biological definition of Alzheimer’s disease. Alzheimer’s & dementia 2018;14:535–562.

21. Jack Jr CR, Andrews JS, Beach TG, et al. Revised criteria for diagnosis and staging of Alzheimer’s disease: Alzheimer’s Association Workgroup. Alzheimer’s & Dementia 2024.

22. Jonaitis EM, Koscik RL, La Rue A, Johnson SC, Hermann BP, Sager MA. Aging, practice effects, and genetic risk in the Wisconsin Registry for Alzheimer’s Prevention. The Clinical Neuropsychologist 2015;29:426–441.

23. Betthauser TJ, Koscik RL, Jonaitis EM, et al. Amyloid and tau imaging biomarkers explain cognitive decline from late middle-age. Brain 2020;143:320–335.

24. Eckerström M, Göthlin M, Rolstad S, et al. Longitudinal evaluation of criteria for subjective cognitive decline and preclinical Alzheimer’s disease in a memory clinic sample. Alzheimer’s & Dementia: Diagnosis, Assessment & Disease Monitoring 2017;8:96–107.

25. Schrag A, Siddiqui UF, Anastasiou Z, Weintraub D, Schott JM. Clinical variables and biomarkers in prediction of cognitive impairment in patients with newly diagnosed Parkinson’s disease: a cohort study. The Lancet Neurology 2017;16:66–75.

26. Gavett BE, Gurnani AS, Saurman JL, et al. Practice Effects on Story Memory and List Learning Tests in the Neuropsychological Assessment of Older Adults. Plos One 2016;11:e0164492.

27. Sanderson Cimino M, Elman JA, Tu XM, et al. Cognitive practice effects delay diagnosis of MCI: Implications for clinical trials. Alzheimer’s & Dementia: Translational Research & Clinical Interventions 2022;8:e12228.

28. Farrell ME, Papp KV, Buckley RF, et al. Association of Emerging β-Amyloid and Tau Pathology With Early Cognitive Changes in Clinically Normal Older Adults. Neurology 2022;98:e1512–e1524.

29. Lim YY, Baker JE, Bruns L, Jr., et al. Association of deficits in short-term learning and Aβ and hippocampal volume in cognitively normal adults. Neurology 2020;95:e2577–e2585.

30. Bennett DA, Schneider JA, Wilson RS, Bienias JL, Arnold SE. Neurofibrillary tangles mediate the association of amyloid load with clinical Alzheimer disease and level of cognitive function. Arch Neurol 2004;61:378–384.

31. Burke DM, Locantore JK, Austin AA, Chae B. Cherry Pit Primes Brad Pitt. Psychological Science 2004;15:164–170.

32. Cohen G. Why is it difficult to put names to faces? British Journal of Psychology 1990;81:287–297.

33. Semenza C, Zettin M. Generating proper names: A case of selective inability. Cognitive neuropsychology 1988;5:711–721.

34. Ramscar M, Hendrix P, Shaoul C, Milin P, Baayen H. The myth of cognitive decline: Non linear dynamics of lifelong learning. Topics in cognitive science 2014;6:5–42.

35. Tennant VR, Harrison TM, Adams JN, Joie RL, Winer JR. Fusiform Gyrus Phospho Tau Is Associated With Failure of Proper Name Retrieval in Aging. Annals of Neurology 2021;90:988–993.

36. Brédart S. The Cognitive Psychology and Neuroscience of Naming People. Neuroscience & Biobehavioral Reviews 2017;83:145–154.

