## Supplemental Materials for "Proper Name Recall as an Early Indicator of Preclinical Alzheimer’s Disease Pathology"

| **Supplemental Table 1**. Total Score Delayed Recall Logical Memory predicting Aβ_42/40_ and p-Tau Status in Nested Models \| **Aβ_42/40_ +/-** \| \| \| **Aβ_42/40_ +/-** \| \| \| **Aβ_42/40_ +/-** \| \|; *Predictors* \| *Odds Ratios* \| *CI* \| *p* \| *Odds Ratios* \| *CI* \| *p* \| *Odds Ratios* \| *CI* \| *p*; Baseline Total Delayed \| 0.9 \| 0.9 – 1.0 \| **0.029** \| 0.9 \| 0.8 – 1.0 \| **0.018** \| 0.9 \| 0.9 – 1.0 \| 0.084; Age at baseline \| \| \| \| 1.1 \| 1.0 – 1.1 \| **<0.001** \| 1.1 \| 1.0 – 1.1 \| **<0.001**; Sex [F] \| \| \| \| 1.1 \| 0.6 – 2.0 \| 0.754 \| 1.0 \| 0.5 – 1.9 \| 0.985; Education \| \| \| \| 1.0 \| 0.9 – 1.1 \| 0.848 \| 1.0 \| 0.9 – 1.1 \| 0.612; Months between last CSF and baseline \| \| \| \| 1.0 \| 1.0 – 1.0 \| **0.006** \| 1.0 \| 1.0 – 1.0 \| **0.018**; *APOE-ε4* [+] \| \| \| \| \| \| \| 7.1 \| 3.8 – 13.7 \| **<0.001**; R2Tjur AIC \| 0.02 319.5 \| \| \| 0.11 302.9 \| \| \| 0.27 263.5 \| \| | | | | | | | | | |
| --- | --- | --- | --- | --- | --- | --- | --- | --- | --- |
|  | **pTau_181_ +/-** | | | **pTau_181_ +/-** | | | **pTau_181_ +/-** | | |
| Baseline Total Delayed | 0.9 | 0.9 – 1.0 | **0.042** | 0.9 | 0.8 – 1.0 | **0.034** | 0.9 | 0.9 – 1.0 | 0.118 |
| Age at baseline |  |  |  | 1.1 | 1.1 – 1.1 | **<0.001** | 1.1 | 1.1 – 1.2 | **<0.001** |
| Sex [F] |  |  |  | 1.1 | 0.6 – 2.1 | 0.805 | 1.0 | 0.5 – 2.1 | 0.932 |
| Education |  |  |  | 0.9 | 0.8 – 1.1 | 0.327 | 0.9 | 0.8 – 1.0 | 0.147 |
| Months between last CSF and baseline |  |  |  | 1.0 | 1.0 – 1.0 | **0.005** | 1.0 | 1.0 – 1.0 | **0.011** |
| *APOE-ε4* [+] |  |  |  |  |  |  | 5.6 | 2.9 – 11.3 | **<0.001** |
| R2Tjur  AIC | 0.02  275.5 |  |  | 0.13  254.8 |  |  | 0.24  230.4 |  |  |

**Model 1** = total score only; **Model 2** = total score + age, sex, education; **Model 3**: Model 2 + *APOE-ε4.* Cut points for CSF amyloid and tau positivity were determined as follows: Aβ_40_/ Aβ_42_ ≤ 0.070, and p-tau_181_ (ptau181 ≥ 51.0 ). CSF values were derived from most recent available visit. Total score delayed recall range: 0-25. *R2 Tjur is computed as the difference in the mean predicted probabilities between the two outcome classes.*

**Supplemental Table 2**. Baseline proper name immediate recall predicting amyloid (top) and pTau_181_ (bottom) status in nested models

|  | **A +/-** | | | **A +/-** | | | **A +/-** | | | |
| --- | --- | --- | --- | --- | --- | --- | --- | --- | --- | --- |
| *Predictors* | *Odds Ratios* | *CI* | *p* | *Odds Ratios* | *CI* | *p* | *Odds Ratios* | | *CI* | *p* |
| Baseline proper name immediate recall | 0.9 | 0.6 – 1.3 | 0.511 | 0.9 | 0.6 – 1.3 | 0.501 | 1.0 | | 0.7 – 1.5 | 0.933 |
| Age at baseline |  |  |  | 1.1 | 1.0 – 1.1 | **<0.001** | 1.1 | | 1.0 – 1.1 | **<0.001** |
| Sex (Female) |  |  |  | 1.0 | 0.5 – 1.7 | 0.885 | 0.9 | | 0.5 – 1.6 | 0.669 |
| Years of Education |  |  |  | 1.0 | 0.9 – 1.1 | 0.903 | 1.0 | | 0.8 – 1.1 | 0.448 |
| Time between baseline and last CSF |  |  |  | 1.0 | 1.0 – 1.0 | **0.008** | 1.0 | | 1.0 – 1.0 | **0.027** |
| *APOE-ε4 (*Carriers) |  |  |  |  |  |  | 7.4 | | 4.0 – 14.3 | **<0.001** |
|  | **T +/-** | | | **T +/-** | | | **T +/-** | | | |
| Baseline proper name immediate recall | 0.9 | 0.6 – 1.3 | 0.495 | 0.9 | 0.6 – 1.3 | 0.495 | 0.9 | 0.6 – 1.3 | | 0.495 |
| Age at baseline |  |  |  | 1.1 | 1.1 – 1.1 | **<0.001** | 1.1 | 0.0 – 0.1 | | **0.002** |
| Sex (Female) |  |  |  | 1.0 | 0.5 – 1.8 | 0.882 | 0.9 | 0.6 – 1.5 | | 0.789 |
| Years of Education |  |  |  | 0.9 | 0.8 – 1.0 | 0.211 | 0.9 | 1.1 – 1.2 | | **<0.001** |
| Time between baseline and last CSF |  |  |  | 1.0 | 1.0 – 1.0 | **0.006** | 1.0 | 0.5 – 1.8 | | 0.789 |
| *APOE-ε4 (*Carriers) |  |  |  |  |  |  | 5.8 | 0.8 – 1.0 | | 0.096 |

**Model 1** = proper names only; **Model 2** = proper names + age, sex, education; **Model 3**: Model 2 + *APOE-ε4.* Cut points for CSF amyloid and tau positivity were determined as follows: Aβ_40_/ Aβ_42_ ≤ 0.070, and pTau181 (pTau181 ≥ 51.0 ). CSF values were derived from most recent available visit. Proper name immediate recall range: 0-4.

**Supplemental Table 3**. Baseline total score immediate recall predicting amyloid (top) and pTau_181_(bottom) status in nested models

|  | **A +/-** | | | **A +/-** | | | **A +/-** | | | |
| --- | --- | --- | --- | --- | --- | --- | --- | --- | --- | --- |
| *Predictors* | *Odds Ratios* | *CI* | *p* | *Odds Ratios* | *CI* | *p* | *Odds Ratios* | | *CI* | *p* |
| Baseline total score immediate recall | 0.9 | 0.6 – 1.3 | 0.511 | 0.9 | 0.6 – 1.3 | 0.501 | 1.0 | | 0.7 – 1.5 | 0.933 |
| Age at baseline |  |  |  | 1.1 | 1.0 – 1.1 | **<0.001** | 1.1 | | 1.0 – 1.1 | **<0.001** |
| Sex (Female) |  |  |  | 1.0 | 0.5 – 1.7 | 0.885 | 0.9 | | 0.5 – 1.6 | 0.669 |
| Years of Education |  |  |  | 1.0 | 0.9 – 1.1 | 0.903 | 1.0 | | 0.8 – 1.1 | 0.448 |
| Time between baseline and last CSF |  |  |  | 1.0 | 1.0 – 1.0 | **0.008** | 1.0 | | 1.0 – 1.0 | **0.027** |
| *APOE-ε4 (*Carriers) |  |  |  |  |  |  | 7.4 | | 4.0 – 14.3 | **<0.001** |
|  | **T +/-** | | | **T +/-** | | | **T +/-** | | | |
| Baseline total score immediate |  | 0.9 | 0.8 – 1.0 | 0.078 | 0.9 | 0.8 – 1.0 | 0.165 | 1.0 | | 0.9 – 1.1 |
| Age at baseline |  |  |  |  | 1.1 | 1.1 – 1.1 | **<0.001** | 1.1 | | 1.1 – 1.2 |
| Sex (Female) |  |  |  |  | 1.0 | 0.5 – 1.9 | 0.999 | 0.9 | | 0.5 – 1.9 |
| Years of Education |  |  |  |  | 0.9 | 0.8 – 1.1 | 0.271 | 0.9 | | 0.8 – 1.0 |
| Time between baseline and last CSF |  |  |  |  | 1.0 | 1.0 – 1.0 | **0.006** | 1.0 | | 1.0 – 1.0 |
| *APOE-ε4 (*Carriers) |  |  |  |  |  |  |  | 5.7 | | 2.9 – 11.5 |

**Model 1** = total score only; **Model 2** = total score + age, sex, education; **Model 3**: Model 2 + *APOE-ε4.* Cut points for CSF amyloid and tau positivity were determined as follows: Aβ_40_/ Aβ_42_ ≤ 0.070, and pTau_181_ (pTau_181_ ≥ 51.0 ). CSF values were derived from most recent available visit. Total score immediate recall range: 0-25.

**Supplemental Table 4.** Association between last A+/- status or T+/- status and longitudinal Logical Memory proper name trajectories, for delayed and immediate recall

|  | **Proper Names Delayed** | | | **Proper Names Immediate** | | | | **Proper Names Delayed** | | | **Proper Names Immediate** | | |
| --- | --- | --- | --- | --- | --- | --- | --- | --- | --- | --- | --- | --- | --- |
| *Predictors* | *Estimates* | *CI* | *p* | *Estimates* | | *CI* | *p* | *Estimates* | *CI* | *p* | *Estimates* | *CI* | *p* |
| Age (centered) | -0.0068 | -0.0153 – 0.0017 | 0.117 | -0.0096 | | -0.018 – -0.001 | **0.029** | -0.0013 | -.001 – .007 | 0.751 | -0.0087 | -0.0172 – -0.0003 | **0.043** |
| Age^2^ | -0.0007 | -0.0011 – -0.0004 | **<0.001** | -0.0005 | | -0.001– -0.0001 | **0.010** | -0.0006 | -.001 – -.0002 | **0.002** | -0.0004 | -0.0008 – -0.0001 | **0.019** |
| Sex (female) | 0.2266 | 0.0669 – 0.3862 | **0.005** | 0.1844 | | 0.034 – 0.335 | **0.017** | 0.2136 | 0.055 – 0.372 | **0.008** | 0.1781 | 0.0274 – 0.3289 | **0.021** |
| Education | 0.0333 | 0.0009 – 0.0656 | **0.044** | 0.0087 | | -0.022 – 0.039 | 0.574 | 0.0296 | -0.002 – 0.062 | 0.073 | 0.0081 | -0.0224 – 0.0386 | 0.603 |
| Practice | 0.0039 | 0.0028 – 0.005 | **<0.001** | 0.0004 | | -0.0001 – 0.002 | 0.479 | 0.0039 | 0.0028 – 0.005 | **<0.001** | 0.0004 | -0.0007 – 0.002 | 0.444 |
| Age at CSF | 0.0252 | 0.0145 – 0.0359 | **<0.001** | 0.0090 | | -0.001– 0.019 | 0.083 | 0.0277 | 0.0169 – 0.0384 | **<0.001** | 0.0098 | -0.0005 – 0.02 | 0.061 |
| A status [+] | -0.139 | -0.3044 – 0.0264 | 0.099 | 0.0710 | | -0.088 – 0.231 | 0.383 |  |  |  |  |  |  |
| Age × A [+] | 0.0052 | -0.0069 – 0.0172 | 0.402 | -0.0023 | | -0.0147 – 0.010 | 0.711 |  |  |  |  |  |  |
| Age^2^× A [+] | -0.0011 | -0.0019 – -0.0004 | **0.004** | -0.0016 | | -0.002 – -0.001 | **<0.001** |  |  |  |  |  |  |
| T status [+] |  |  |  |  | |  |  | -0.129 | -0.303 – 0.044 | 0.144 | 0.0594 | -0.1085 – 0.227 | 0.488 |
| Age × T status [+] |  |  |  |  | |  |  | -0.008 | -0.019 – 0.0048 | 0.228 | -0.0027 | -0.0154 – 0.010 | 0.680 |
| Age^2^ ×  T status [+] |  |  |  |  | |  |  | -0.002 | -0.002 – -0.001 | **<0.001** | -0.0018 | -0.0026 – -0.001 | **<0.001** |
| Conditional R^2^ | 0.507 | | | | 0.389 | | | 0.508 | | | 0.387 | | |

*Proper Names:* Names of people or places from Logical Memory Story A (range = 0-4); Age (years and months) at Logical Memory visits, centered at the mean of 68 years; Age^2^=centered age squared, quadratic term. *Practice:* Maximum visit number of Logical Memory – 1; Aβ_42/40_ Status [A+/A-] and pTau181 Status [T+/T-] based on published cut points^23^ (Aβ_40_/Aβ_42_ ≤ 0.070, pTau181 ≥ 51.0 ). Models are adjusted for random effects of slope and intercept. Significance was set at p<.05.

**Supplemental Table 5.** Association of last A+/- status or T+/- status with longitudinal Logical Memory total score trajectories, separately for delayed and immediate recall

|  | **Total Score Delayed** | | | **Total Score Immediate** | | | **Total Score Delayed** | | | **Total Score Immediate** | | |
| --- | --- | --- | --- | --- | --- | --- | --- | --- | --- | --- | --- | --- |
| *Predictors* | *Estimates* | *CI* | *p* | *Estimates* | *CI* | *p* | *Estimates* | *CI* | *p* | *Estimates* | *CI* | *p* |
| Age | 0.0150 | 0.0050 – 0.0251 | **0.003** | 0.0206 | 0.0110 – 0.0303 | **<0.001** | 0.0175 | 0.0079 – 0.0272 | **<0.001** | 0.0193 | 0.0124 – 0.0263 | **<0.001** |
| Age^2^ | -0.0011 | -0.002 – -0.001 | **<0.001** | -0.0006 | -0.0010 – -0.0002 | **0.003** | -0.001 | -0.0014 – -0.001 | **<0.001** | -0.0001 | -0.0003 – 0.0002 | 0.494 |
| Sex [F] | 0.2859 | 0.1065 – 0.4653 | **0.002** | 0.3334 | 0.1544 – 0.5124 | **<0.001** | 0.2905 | 0.1110 – 0.4700 | **0.002** | 0.3669 | 0.1823 – 0.5515 | **<0.001** |
| Education | 0.0501 | 0.0135 – 0.0866 | **0.007** | 0.0301 | -0.0061 – 0.0663 | 0.103 | 0.0472 | 0.0106 – 0.0838 | **0.011** | 0.0147 | -0.0229 – 0.0523 | 0.443 |
| practice | 0.0024 | 0.0014 – 0.0035 | **<0.001** | -0.0003 | -0.0014 – 0.0007 | 0.553 | 0.0024 | 0.0014 – 0.0035 | **<0.001** | 0.0009 | -0.0001 – 0.0020 | 0.082 |
| Age at CSF | 0.0357 | 0.0237 – 0.0477 | **<0.001** | 0.0299 | 0.0180 – 0.0419 | **<0.001** | 0.0383 | 0.0262 – 0.0504 | **<0.001** | 0.0356 | 0.0232 – 0.0480 | **<0.001** |
| A [+] | -0.2472 | -0.428 – -0.067 | **0.007** | -0.1705 | -0.3526 – 0.0116 | 0.066 |  |  |  |  |  |  |
| Age * A [+] | -0.0047 | -0.0199 – 0.0105 | 0.542 | -0.0134 | -0.0276 – 0.0007 | 0.063 |  |  |  |  |  |  |
| Age^2^ * A [+] | -0.0014 | -0.002 – -0.001 | **0.001** | -0.0020 | -0.0028 – -0.0012 | **<0.001** |  |  |  |  |  |  |
| T [+] |  |  |  |  |  |  | -0.236 | -0.427 – -0.046 | **0.015** | -0.175 | -0.379 – 0.029 | 0.093 |
| Age * T [+] |  |  |  |  |  |  | -0.008 | -0.024 – 0.007 | 0.287 | -0.01 | -0.019 – -0.002 | **0.021** |
| Age^2^ * T [+] |  |  |  |  |  |  | -0.002 | -0.003– -0.001 | **<0.001** | -0.001 | -0.001 – -0.0004 | **<0.001** |
| Conditional R^2^ | 0.501 | | | 0.602 | | | 0.505 | | | 0.599 | | |

*Total Score:* Idea units from Logical Memory Story A (range = 0-25); Age (years and months) at Logical Memory visits, centered at the mean of 68 years; Age^2^=centered age squared, quadratic term. *Practice:* Maximum visit number of Logical Memory – 1; Aβ_42/40_ Status [A+/A-] and pTau_181_ Status [T+/T-] based on published cut points^23^ (Aβ_40_/ Aβ_42_ ≤ 0.070, pTau181 ≥ 51.0 ). Models are adjusted for random effects of slope and intercept. Significance was set at p<.05

Supplemental Table 6. Sensitivity analysis of 3 level A/T status variable predicting delayed and immediate proper name total score from Logical Memory

|  | **Proper Name Delayed** | | | **Proper Name Immediate** | | | **Total Score Delayed** | | | **Total Score Immediate** | | |
| --- | --- | --- | --- | --- | --- | --- | --- | --- | --- | --- | --- | --- |
| *Predictors* | *Estimates* | *CI* | *p* | *Estimates* | *CI* | *p* | *Estimates* | *CI* | *p* | *Estimates* | *CI* | *p* |
| Age (centered) | -0.001 | -0.005 – 0.0022 | 0.500 | -0.002 | -0.004 – 0.000 | 0.054 | 0.0048 | 0.0030 – 0.0066 | **<0.001** | 0.0046 | 0.0031 – 0.0061 | **<0.001** |
| Age^2^ | -0.0002 | -0.0003 – -0.0001 | **0.006** | -0.0001 | -0.0002 – -.000 | **0.033** | -.0001 | -.0001 – 0.0000 | 0.052 | 0.0000 | -.0000 – 0.0001 | 0.733 |
| Sex [F] | 0.0823 | 0.0190 – 0.1456 | **0.011** | 0.0351 | -.0024 – .0725 | 0.067 | 0.0789 | 0.0294 – 0.1284 | **0.002** | 0.0683 | 0.0276 – 0.1091 | **0.001** |
| Education | 0.0106 | -0.0021 – 0.0232 | 0.101 | 0.0022 | -.0052 – .0096 | 0.562 | 0.0074 | -.0025 – .0173 | 0.143 | 0.0038 | -.0044 – .0119 | 0.368 |
| Practice | 0.0015 | 0.0011 – 0.0020 | **<0.001** | 0.0000 | -.0002 – .0003 | 0.852 | 0.0010 | 0.0007 – 0.0012 | **<0.001** | 0.0002 | -.0001 – .0004 | 0.143 |
| CSF Age | 0.0009 | 0.0005 – 0.0013 | **<0.001** | 0.0002 | -.0000 – 0.0004 | 0.084 | 0.0008 | 0.0005 – 0.0010 | **<0.001** | 0.0006 | 0.0003 – 0.0008 | **<0.001** |
| A/T Status [A+/T-] | -0.0135 | -0.1074 – 0.0804 | 0.778 | 0.0102 | -.0462 – 0.0667 | 0.722 | -.0262 | -.1005 – 0.0480 | 0.489 | -.0084 | -.0699 – 0.0530 | 0.788 |
| A/T Status [A+/T+] | -0.0813 | -0.1575 – -0.0050 | **0.037** | 0.0167 | -.0291 – 0.0625 | 0.474 | -.1006 | -.1600 – -.0411 | **0.001** | -.0590 | -.1083 – -.0098 | **0.019** |
| Age × A/T Status [A+/T-] | 0.0046 | -0.0020 – 0.0112 | 0.170 | 0.0021 | -.0023 – 0.0065 | 0.354 | -.0012 | -.0041 – 0.0016 | 0.400 | -.0022 | -.0047 – 0.0003 | 0.081 |
| Age × A/T Status [A+/T+] | -0.0008 | -0.0062 – 0.0046 | 0.771 | -0.0013 | -.0048 – 0.0022 | 0.472 | -.0020 | -.0044 – 0.0004 | 0.097 | -.0031 | -.0052 – -.0009 | **0.005** |
| Age^2^ × A/T Status [A+/T-] | -0.0003 | -0.0008 – 0.0002 | 0.186 | -0.0002 | -.0005 – 0.0001 | 0.283 | -.0004 | -.0006 – -.0001 | **0.001** | -.0005 | -.0007 – -.0003 | **<0.001** |
| Age^2^ × A/T Status [A+/T+] | -0.0005 | -0.0009 – -0.0002 | **0.001** | -0.0005 | -.0007 – -.0002 | **<0.001** | -.0005 | -.0006 – -.0003 | **<0.001** | -.0005 | -.0006 – -.0004 | **<0.001** |
| Marginal R^2^ / Conditional R^2^ | 0.138 / 0.485 | | | 0.054 / 0.399 | | | 0.224 / 0.666 | | | 0.161 / 0.603 | | |

*Proper Names:* Names of people or places from Logical Memory Story A (range = 0-4); *Total Score:* Idea units from Logical Memory Story A (range = 0-25); Age (years and months) at Logical Memory visits, centered at the mean of 68 years; Age^2^=centered age squared, quadratic term. *Practice:* Maximum visit number of Logical Memory – 1; Aβ_42/40_ Status [A+/A-] and pTau181 Status [T+/T-] based on published cut points^23^ (Aβ_40_/ Aβ_42_ ≤ 0.070, pTau181 ≥ 51.0 ). *AT sttaus:* Amyloid/Tau status was a 3-level variable, where those who were tau positive/amyloid negative were excluded from the study sample. Models are adjusted for random effects of slope and intercept. Significance was set at p < .05

| **Supplemental Table 7.** Linear mixed effects model results showing three-way interaction among practice, age, and biomarker status predicting Logical Memory outcomes | | | | | | | | | | | | |
| --- | --- | --- | --- | --- | --- | --- | --- | --- | --- | --- | --- | --- |
|  | **Proper Names Delayed** | | | **Total Score Delayed** | | | **Proper Names Delayed** | | | **Total Score Delayed** | | |
| *Predictors* | *Estimates* | *CI* | *p* | *Estimates* | *CI* | *p* | *Estimates* | *CI* | *p* | *Estimates* | *CI* | *p* |
| Age | 0.008 | 0.004 – 0.011 | **<0.001** | 0.031 | 0.025 – 0.037 | **<0.001** | 0.007 | 0.005 – 0.010 | **<0.001** | 0.033 | 0.027 – 0.038 | **<0.001** |
| Sex [F] | 0.132 | 0.067 – 0.197 | **<0.001** | 0.418 | 0.225 – 0.612 | **<0.001** | 0.132 | 0.065 – 0.198 | **<0.001** | 0.431 | 0.238 – 0.625 | **<0.001** |
| Education | 0.016 | 0.003 – 0.029 | **0.019** | 0.046 | 0.007 – 0.085 | **0.022** | 0.012 | -0.001 – 0.026 | 0.071 | 0.043 | 0.004 – 0.082 | **0.031** |
| Practice | 0.002 | -0.003 – 0.007 | 0.405 | 0.027 | 0.017 – 0.038 | **<0.001** | 0.006 | 0.001 – 0.010 | **0.016** | 0.028 | 0.018 – 0.038 | **<0.001** |
| Age at CSF | 0.001 | 0.001 – 0.001 | **<0.001** | 0.004 | 0.002 – 0.005 | **<0.001** | 0.001 | 0.001 – 0.002 | **<0.001** | 0.004 | 0.003 – 0.005 | **<0.001** |
| Aβ_42/40_ [+] | -0.151 | -0.22– -0.07 | **<0.001** | -0.552 | -.756 – -.347 | **<0.001** |  |  |  |  |  |  |
| Age × Aβ_42/40_ [+] | 0.002 | -0.004 – 0.01 | 0.606 | 0.001 | -0.010 – 0.012 | 0.864 |  |  |  |  |  |  |
| Practice × Aβ_42/40_ [+] | 0.014 | 0.004 – 0.024 | **0.008** | 0.023 | 0.001 – 0.045 | **0.042** |  |  |  |  |  |  |
| Age × practice | -0.001 | -.001 – -.000 | **0.003** | -0.002 | -.003 – -.001 | **<0.001** | -0.000 | -.001 – .000 | 0.067 | -0.001 | -.002 – -0001 | **0.002** |
| Age × practice × Aβ_42/40_ [+] | -0.001 | -.002 – -.000 | **0.009** | -0.003 | -.005 – -.001 | **0.001** |  |  |  |  |  |  |
| pTau [+] |  |  |  |  |  |  | -0.168 | -.245 – -.091 | **<0.001** | -0.558 | -.773 – -.344 | **<0.001** |
| Age × pTau [+] |  |  |  |  |  |  | 0.001 | -.004 – .006 | 0.746 | -0.004 | -.016 – .007 | 0.454 |
| Practice × pTau [+] |  |  |  |  |  |  | 0.014 | 0.003 – 0.025 | **0.013** | 0.028 | 0.004 – 0.052 | **0.023** |
| Age × pTau [+] × Practice |  |  |  |  |  |  | -0.002 | -.003 – -.001 | **<0.001** | -0.004 | -.006 – -.002 | **<0.001** |
| Marginal R^2^ / Conditional R^2^ | 0.105 / 0.521 | | | 0.219 / 0.674 | | | 0.127 / 0.483 | | | 0.220 / 0.676 | | |

*Proper Names:* Names of people or places from Logical Memory Story A (range = 0-4); *Total Score:* Idea units from Logical Memory Story A (range = 0-25); Age (years and months) at Logical Memory visits, centered at the mean of 68 years; Age^2^=centered age squared, quadratic term. *Practice:* Maximum visit number of Logical Memory – 1; Aβ_42/40_ Status [A+/A-] and pTau181 Status [T+/T-] based on published cut points^23^ (Aβ_40_/ Aβ_42_ ≤ 0.070, pTau181 ≥ 51.0 ). Models are adjusted for random effects of slope and intercept. Significance was set at p < .05

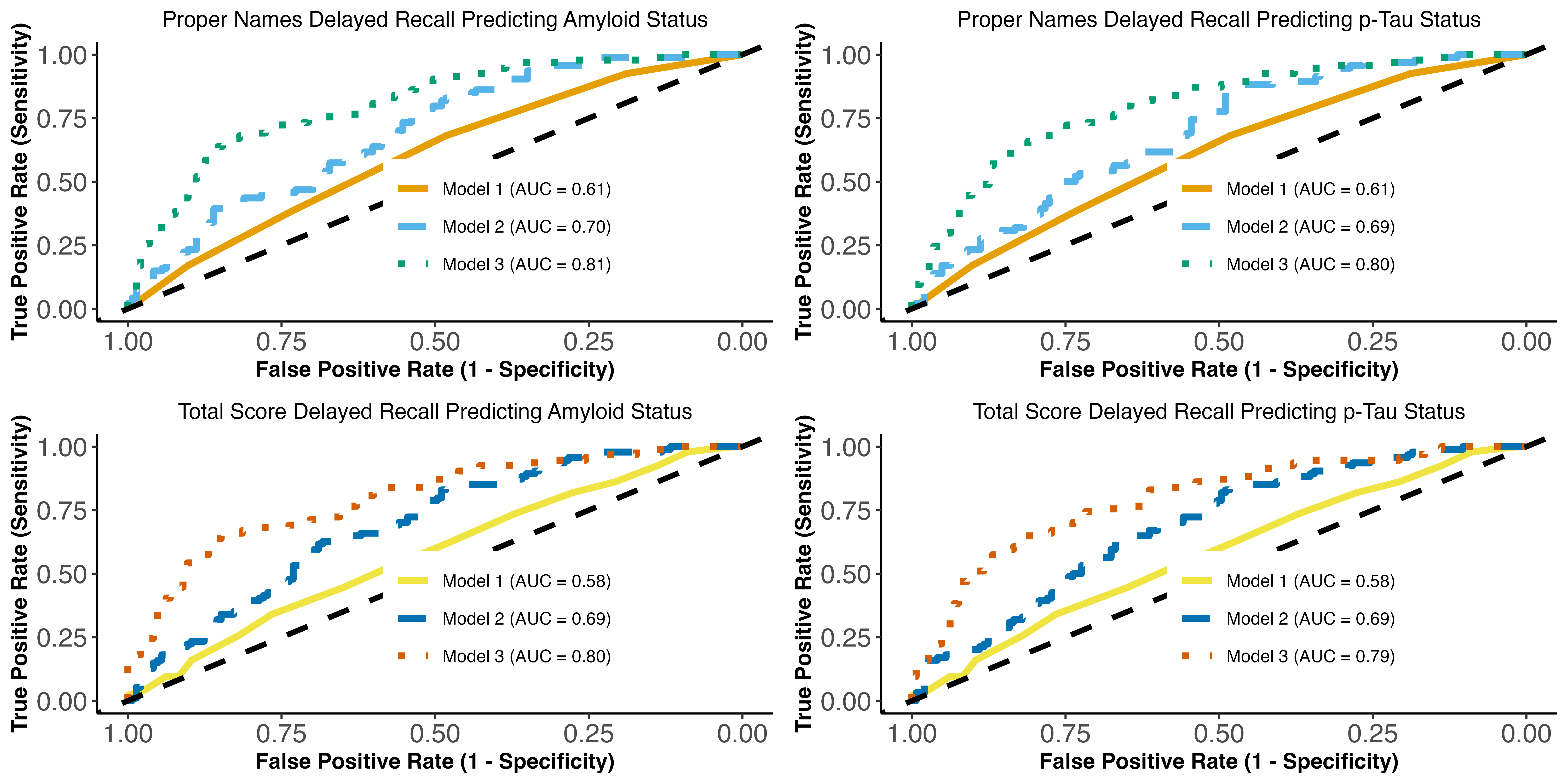

***Supplemental Figure 1:*** *ROC curves for baseline* ***proper names*** *(top) and* ***total score*** *(bottom) delayed recall predicting CSF Aβ_42/40_ (+/-) status (left) and CSF pTau_181_ (+/-) status (right) in nested models*. N = 271 study participants. **Model 1** = proper names only; **Model 2** = proper names + age, sex, education; **Model 3**: Model 2 + *APOE-ε4.* The cut-off points for CSF amyloid and tau positivity were determined as follows: Aβ_40_/ Aβ_42_ ≤ 0.070 and p-tau_181_ (ptau181 ≥ 51.0 ). CSF values were derived from the most recent available visit. Proper name delayed recall range: 0-4. *ROC:* Receiver Operating Curve; *AUC =* Area under the curve.
